# Left Atrial Stiffness Identifies Children with Magnetic-Resonance-Imaging-Proven Acute Myocarditis Despite Preserved Left Ventricular Systolic Function

**DOI:** 10.1101/2025.08.01.25332594

**Authors:** Ryusuke Numata, Renzo Calderon-Anyosa, Danish Vaiyani, Sarina Sun, Yan Wang, Laura Mercer-Rosa, David M. Biko, Anirban Banerjee

## Abstract

**Background:** Left atrium (LA) may be labeled as a “forgotten” chamber in echocardiographic evaluation of pediatric myocarditis. Recently, LA stiffness has gained attention in identifying both diastolic dysfunction and myocardial injury in children.

**Methods:** We retrospectively analyzed 51 pediatric patients with acute myocarditis diagnosed by cardiac MRI based on updated Lake-Louise-Criteria, along with 40 age-matched healthy controls. Only patients with preserved LVEF (>55%) were included, given their increased risk of adverse outcomes due to diastolic impairment. Follow-up imaging was available for 41 of the 51 patients. LV systolic and diastolic function, and LA strain were evaluated by conventional 2D and speckle-tracking echocardiography. LA stiffness was calculated as the ratio of E/e’ to peak LA strain as depicted in following equation:

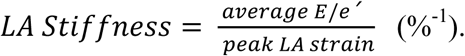

**Results:** LA stiffness was significantly increased in myocarditis patients (0.27 ± 0.09 vs. 0.15 ± 0.04 %^-1^, p<0.001) and remained impaired at early follow-up. LA stiffness showed the best correlation with peak BNP (r = 0.66, p<0.001). Moreover, LA stiffness had the highest diagnostic performance among all echocardiographic indices, with an AUC of 0.94, and remained an independent diagnostic power in multivariable regression model (OR 1.58 [95% CI: 1.32 – 1.89], p<0.001). When it was incorporated into a composite score with LV peak longitudinal strain, the AUC yielded the highest with 98% sensitivity.

**Conclusion:** An optimal LA stiffness cutoff of 0.2%^-1^ may provide incremental value as a new diagnostic marker for the diagnosis of acute myocarditis with preserved LVEF in children, when this value is exceeded.

**Clinical Perspective:** *What Is New?:*

- We evaluated the diagnostic utility of left atrial (LA) stiffness in pediatric patients with acute myocarditis and preserved left ventricular ejection fraction (LVEF), using both conventional and two-dimensional speckle-tracking echocardiography.
- LA stiffness was significantly elevated alongside reduced peak LA strain in pediatric myocarditis patients confirmed by cardiac magnetic resonance imaging (CMR), even during early follow-up, reflecting persistent left ventricular diastolic dysfunction.
- LA stiffness demonstrated the highest diagnostic accuracy in distinguishing patients with acute myocarditis from normal control, with an AUC of 0.94 (cutoff of 0.2%^-1^), and incorporating it with LV longitudinal strain enhanced the diagnostic performance.

*What Are The Clinical Implications?:*

- LA strain analysis may play a pivotal role in helping clinicians identify diastolic dysfunction and detect pediatric patients with CMR-proven acute myocarditis, even with preserved LVEF, facilitating early diagnosis and support tailored management during follow-up.

## Background

Acute myocarditis is a serious illness that may lead to significant morbidity and mortality. A nationwide study in the United States revealed an in-hospital mortality rate of 6.8% among children,^1^ emphasizing the higher risk of pediatric patients compared to adults, who had an overall mortality rate of 2.3%.^2^ The incidence of myocarditis has been increasing from 0.7 to 0.9 per 100,000 in children over the past decade, alongside the growing recognition of the diagnostic role of cardiac magnetic resonance imaging (CMR).^1,3^ Although CMR allows for superior tissue characterization, typically the first imaging test performed in most pediatric patients with suspected myocarditis is an echocardiogram. However, early in the course of myocarditis, left ventricular (LV) dimensions are generally normal, and LV ejection fraction (LVEF) is preserved in approximately 75% of patients, making an echocardiographic diagnosis quite challenging.^4^ Additionally, given the heterogenous clinical presentation of myocarditis, pediatric patients often present with limited symptoms, such as non-specific chest pain, which can lead to the underuse of myocarditis-focused diagnostic tools, like laboratory tests for troponin levels or CMR.^5^ For logistic reasons and lack of resources it may not be possible to obtain a CMR during early presentation of chest pain. Consequently, the incidence of pediatric myocarditis may be underestimated.^6^ Despite the absence of a singular definitive diagnostic test, cardiac function may decline rapidly during the initial days after presentation.^2^ Moreover, myocarditis patients with diastolic impairment, including those with normal-EF, have an increased risk of adverse clinical outcomes during follow-up. Specifically, acute myocarditis with preserved-EF has been associated with a higher hospitalization rate over a 5-year follow-up, attributed to diastolic dysfunction. This underscores the critical role of diastolic functional assessment using echocardiography.^7^ Therefore, comprehensive echocardiographic assessment that incorporates an index with focused diagnostic accuracy into the standard routine protocol, is essential for both initial diagnosis and follow-up management of acute myocarditis patients, even when LVEF is preserved.

In recent years, left atrial (LA) strain analysis using two-dimensional speckle-tracking echocardiography (2D-STE) has emerged as a novel imaging method with superior diagnostic value for patients with heart failure with preserved-EF (HFpEF).^8^ Since the LA and LV are intimately connected during ventricular diastole, it is intuitive to examine the physiologic features of the LA in patients with impaired diastolic function. Several studies have shown that LA functional analysis can offer more sensitive and accurate assessment of diastolic impairment compared with traditional diastolic indices, even in the early stages of cardiac diseases.^9,10^ More recently, LA stiffness obtained by 2D-STE has demonstrated greater diagnostic and even prognostic power than LA strain alone in adult patients with HFpEF according to the latest guideline for the evaluation of left ventricular diastolic function recently updated by the American Society of Echocardiography (ASE).^11,12^ Our previous study in children with multisystem inflammatory syndrome associated with COVID-19 has highlighted the advantages of evaluating noninvasive LA stiffness, which serves as a surrogate marker to differentiate elevated pulmonary capillary wedge pressure (PCWP) from normal and to predict COVID-19 related myocardial injury.^13^ In our recent study in children with hypertrophic cardiomyopathy (HCM), LA stiffness showed the strongest prognostic value for predicting adverse cardiac events, outperforming all other echocardiographic parameters, including conventional risk factors. Together, these findings from our previous work highlight the superiority of LA stiffness in detecting diastolic impairment even in the pediatric population.^14^

In acute myocarditis, adult patients with preserved EF exhibit impaired LA strain and stiffness, even during follow-up.^9,15^ Although some reports on myocarditis have referred to the diagnostic and prognostic value of strain analysis in both LV and LA, these assessments were primarily conducted using CMR and were limited to adult cohorts.^16,17^ Moreover, longitudinal 2D-STE evaluation can be of incremental value in monitoring ongoing LV diastolic dysfunction.^16^

Therefore, the aim of this study is to evaluate the enhanced diagnostic value of LA stiffness and to assess longitudinal LA deformation using 2D-STE in pediatric myocarditis patients with preserved-EF. We hypothesize that LA stiffness will demonstrate superior diagnostic performance in identifying pediatric patients with acute myocarditis, despite preserved-EF. We further propose that longitudinal deformation analysis of the LA will reveal impaired LA strain and increase LA stiffness from the acute phase to early follow-up.

## Methods

### Study design

This is a retrospective, longitudinal study evaluated at The Children’s Hospital of Philadelphia between 2011 to 2024. We included 111 pediatric patients under 21 years of age, all of whom underwent CMR and were diagnosed with acute myocarditis based on the updated Lake Louise Criteria (LLC).^3^ Inclusion criteria required the availability of a transthoracic echocardiogram (TTE) at the time of diagnosis confirmed by CMR. As some patients had undergone echocardiography at referring institutions, which were not accessible for subsequent analysis, they were excluded. An LVEF greater than 55% was also required for inclusion in this study. For the early follow-up cohort, we included the most recent echocardiographic study after discharge. CMR was conducted in the acute phase of myocarditis within one week of the first evaluation of the patients, suspected of acute myocarditis based on clinical manifestations, laboratory findings, and electrocardiographic (ECG) alterations. Exclusion criteria included viral myocarditis patients with proven COVID-19 related myocardial injury, inadequate imaging quality, and inability to access TTE data. Fifty-one patients were included in the acute myocarditis cohort and forty-one patients were enrolled in the early follow-up cohort. Additionally, forty and thirty-nine healthy age-matched children served as controls for acute and follow-up cohort, respectively. These controls were selected from children with structurally normal hearts who underwent echocardiography for the evaluation of benign heart murmurs, chest pain, syncope, or a family history of cardiac disease. In the acute myocarditis cohort, we assessed the diagnostic performance of 2D-STE and conventional indices for detecting acute myocarditis. Laboratory test results, such as peak brain natriuretic peptide (BNP) and peak troponin I (TnI) levels, were obtained from the patient-records. The Institutional Review Board of the Children’s Hospital of Philadelphia approved the study, with a waiver of informed consent granted.

### Echocardiography

All echocardiographic studies were performed using iE33 or EPIC CVx (Phillips Medical Systems, Andover, MA, USA). Conventional echocardiographic measurements were obtained according to the ASE guidelines^12,18^, including standard LV diameters in the parasternal short-axis view by M-mode, LVEF using Simpson’s biplane method. As per the convention in our echocardiography laboratory, LA volumes were not available; instead, antero-posterior dimensions of the LA were measured from parasternal long-axis images when the LV was at end systole. These LA dimensions were converted to z scores using the Boston z-score calculator.^19^ Additionally, isovolumic relaxation time (IVRT) between aortic valve closure and mitral valve opening was measured from corresponding Doppler signals.

### Speckle-tracking echocardiography

The digital imaging and communications in medicine (DICOM) clips of the 2D apical four-chamber view of the LA, were uploaded to a vendor-independent STE software (2D Cardiac Performance Analysis 1.3.0.91; TomTec Imaging Systems, Munich, Germany) to trace the LA endocardial border. The LA tracing for the strain analysis was terminated 0.3 cm above the atrio-ventricular junction, to minimize the effect of LV myocardial movement. The software generated average peak positive LA strain values. Visual tracking of the LA wall was performed on the cine images. To be consistent with previous studies, the LV end-diastole at the onset of the QRS complex was defined as the starting point of the atrial cardiac cycle (Figure 2A).

We also calculated the LV peak longitudinal strain (LVLS) by tracing the LV endocardial border using the same software. Due to lack of adequate 3- and 2-chamber views in early years of our study, those were not included. In addition, longitudinal early diastolic strain rate (EDSR_L_) was measured from the same image of the LV.^20^

### Measure of LA stiffness

As the stiffness of a biological material is described by its stress-strain relationship, Kurt *et al.* proposed an index for the stiffness of the LA as the ratio of the PCWP to the LA strain.^8^ Using the E/e’ ratio as a surrogate of PCWP in adults, they defined the noninvasive LA stiffness index as the ratio of E/e’ to LA strain as depicted in the following equation:

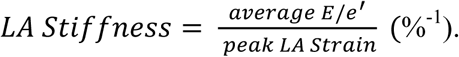

LA stiffness has demonstrated a distinct cutoff value in children for detecting LV filling pressures of >12 mmHg with high degree of accuracy.^13^

### Cardiac magnetic resonance imaging

CMR was performed within 7 days of presentation using a 1.5- or 3-Tesla Siemens MRI system (Siemens Medical Solutions, Erlangen, Germany) with a 32-channel phased-array receiver coil, per the clinical protocol at our institution. Post-processing was conducted using cvi42 software version 5.12.4 (Circle Cardiovascular Imaging, Calgary, Canada). The protocol for patients with suspected myocarditis included coronary angiography using inversion recovery gradient echo imaging with ECG gating and navigators for respiratory motion adaptation. It also involved short-axis projection views consisting of Modified Look-Locker Inversion recovery sequences for T1 mapping, T2-weighted turbo-inversion recovery magnitude sequences to assess signal intensity ratios of cardiac and skeletal muscle, and T2-prepared single-shot steady-state-free-precession sequences for T2 mapping. Late gadolinium enhancement assessment was performed in the 4-chamber, long-axis, and short-axis views using respiratory motion-corrected magnitude and phase-sensitive inversion recovery sequences. CMR studies showing both myocardial edema (T2-based criterion) and non-ischemic myocardial injury (T1-based criterion) are required for the diagnosis of acute myocarditis, following the ‘2 out of 2’ approach according to updated LLC.^3^

### Intra- and Inter-observer Variability

Intra- and inter-observer reliability of LA strain and LA stiffness measurements were assessed using Bland–Altman analysis and intraclass correlation coefficients (ICCs) with bias and 95% limits of agreement (LOA). For intra-observer variability, repeated measurements were performed by the same observer on 30 randomly selected patients more than 2 weeks apart, using ICC (3,1) (two-way mixed-effects model, single measures) to assess consistency. Inter-observer variability was evaluated by two independent, blinded observers analyzing LA function offline in another set of 30 randomly selected subjects, using ICC (2,1) (two-way random-effects model, single measures) to assess agreement.

### Statistical analysis

Continuous variables were expressed as mean ± SD or median (25th, 75th percentiles), while categorical variables were displayed as percentages. Baseline characteristics were compared using Fisher’s exact tests for categorical variables and Student’s *t* test for continuous variables, as appropriate. Echocardiographic measurements between the two independent groups were assessed using two-sample Student’s *t* tests or Mann-Whitney U tests, as appropriate. Differences between continuous variables in paired data were assessed with a paired *t*-test. Echocardiographic parameters with significant differences were further examined using receiver operating characteristic (ROC) curve analysis to determine their area under the curve (AUC) and cut-off value for establishment of the optimal diagnostic threshold based on the maximum Youden index. AUC were compared using the DeLong test.^21^

Correlations among LA stiffness, LVLS, BNP, and TnI were calculated using Pearson’s correlation coefficients, and differences between correlations were assessed for significance. Multivariable logistic regression was used to evaluate the independent effect of each variable on the presence of myocarditis. We combined LA stiffness and LVLS into a composite score, as both demonstrated significant predictive performance in identifying acute myocarditis with preserved-LVEF, without collinearity. However, the coefficients derived from multivariable analysis used to construct this equation resulted in a cumbersome formula, which could hinder clinical acceptability. To address this, we have simplified the equation by using a straightforward subtraction of the absolute values after confirming the discriminative values were equivalent in each model derived from ROC analysis. Due to small, decimal values of LA stiffness, the value was multiplied by 100 in this model to represent the change in LA stiffness for each decimal point increase from 0.01 to 0.02. This adjustment was also applied to the multivariable logistic regression analysis. The composite score was further analyzed using ROC analysis to determine the optimal cut-off value.

All analyses were performed using R version 4.4.2. Measurements were considered reproducible if ICCs were > 0.75. For all analyses, a P-value of less than 0.05 was considered statistically significant.

## Results

### Acute myocarditis cohort

#### Demographic data

In this cohort, there were fifty-one patients had preserved-LVEF and met inclusion criteria (Figure 1). The acute myocarditis group had significantly larger BSA and BMI, despite being age- and sex-matched to the controls. (Table 1) The most common symptom was chest pain, which was present in 92.2% of myocarditis patients, accompanied by elevated TnI levels in all patients in our cohort and moderately elevated BNP values. Although the causative viruses for presumed viral myocarditis were often undetected by laboratory testing, nearly half of the patients showed preceding signs of infection, with 64.3% presenting with upper respiratory symptoms and 35.7 % showing gastrointestinal symptoms. ECG abnormalities were found in roughly three-fourths of the patients, with half showing various ST segment changes. Patients were largely treated with supportive therapy, including non-steroidal anti-inflammatory drugs.

**Figure 1.**
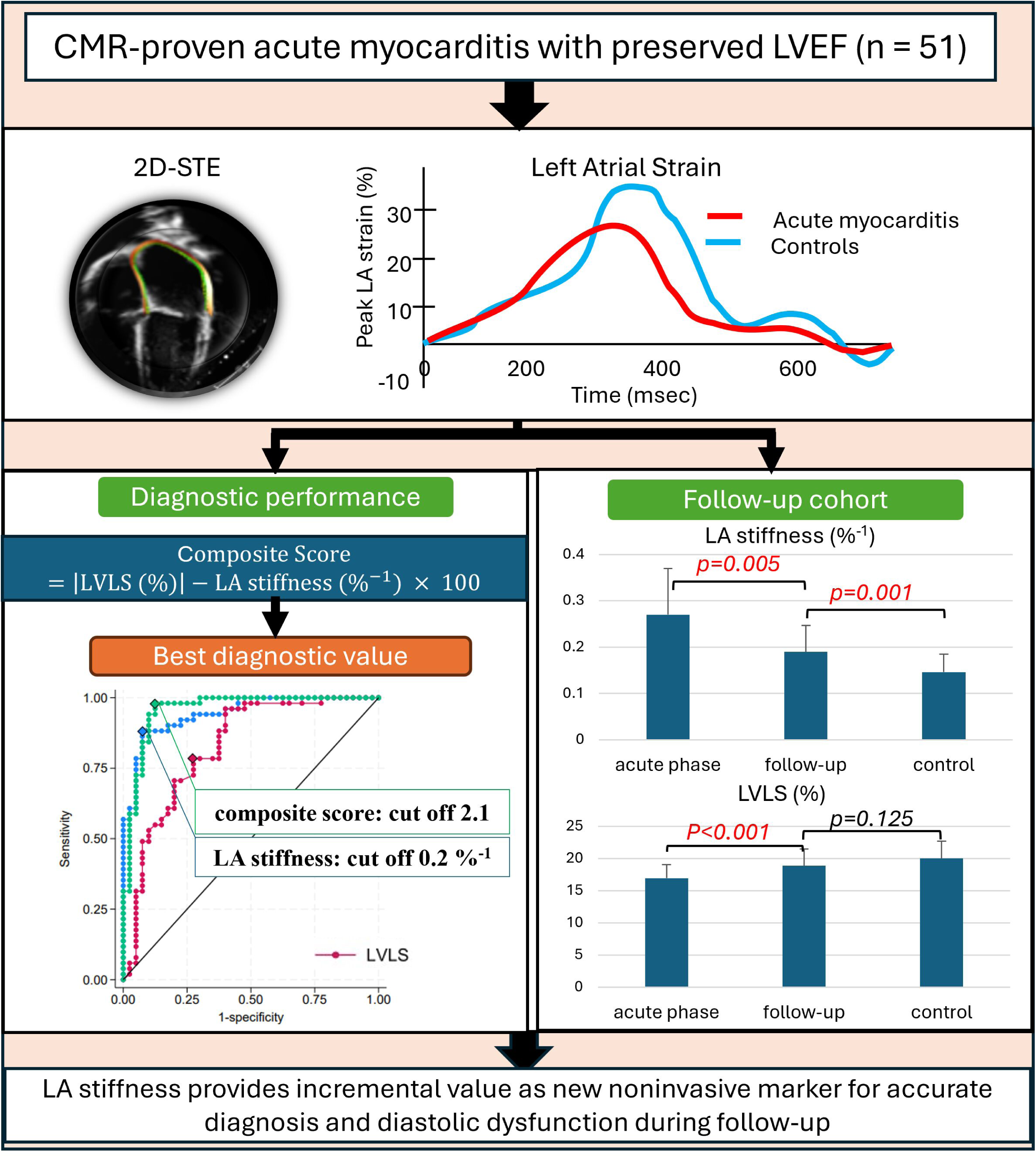
Patient recruitment and selection flowchart. This flowchart details the recruitment, selection, and classification of patients for the study, adhering to the updated Lake Louise Criteria (LLC) for Cardiac Magnetic Resonance in Nonischemic Myocardial Inflammation Guidelines.

**Table 1.**
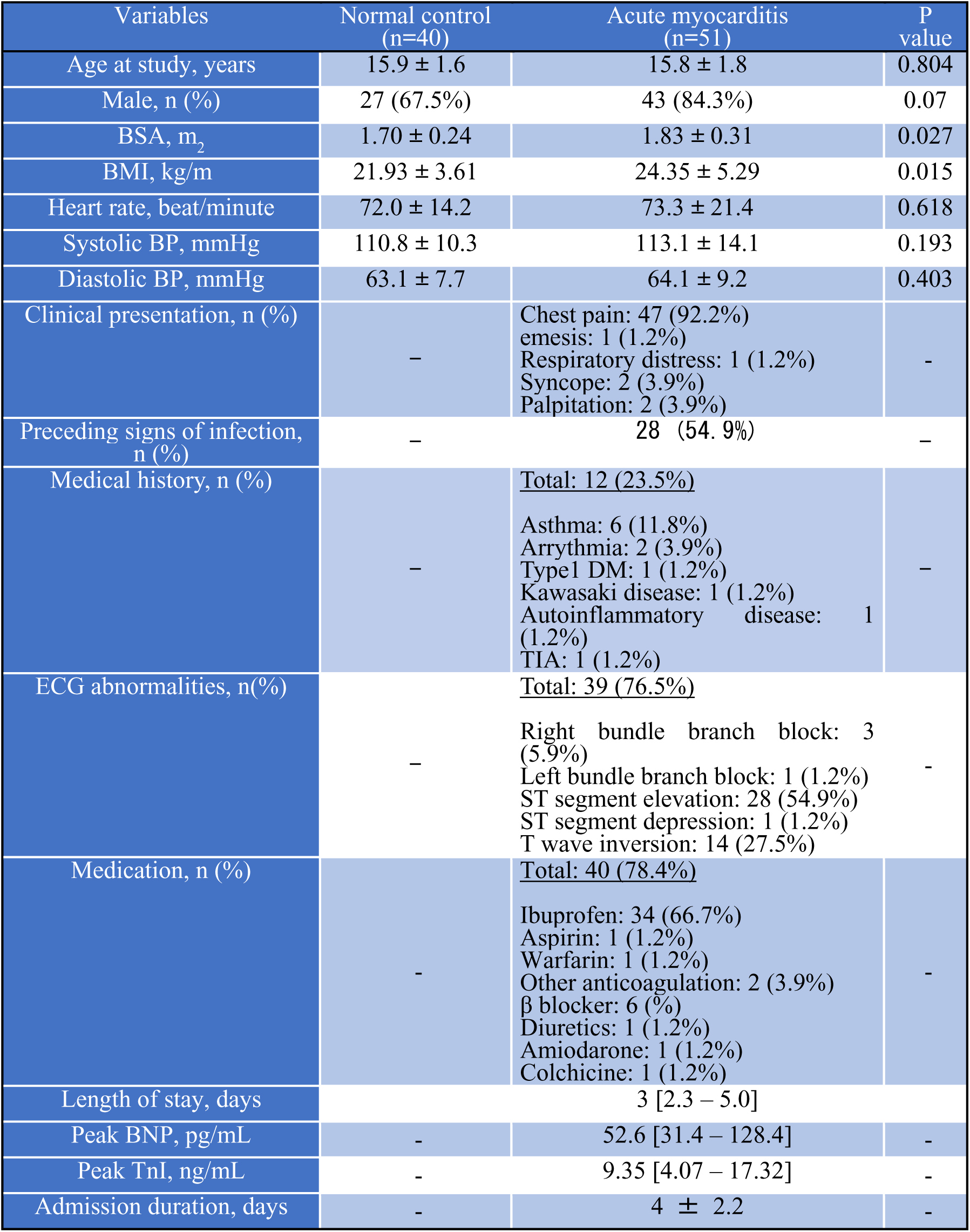
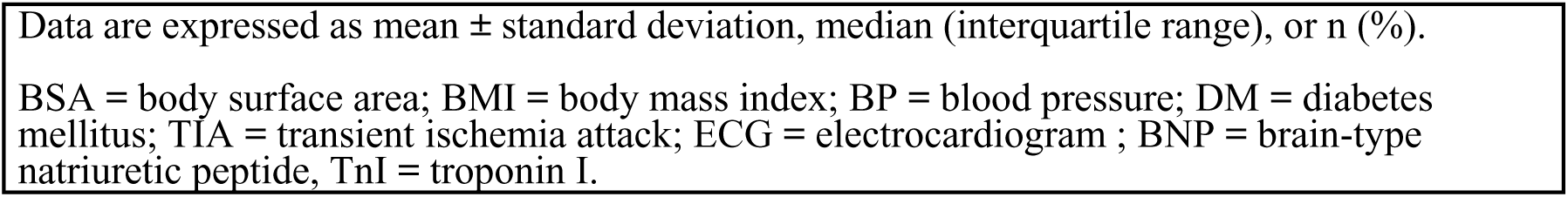
Demographics.

**Table 2.**
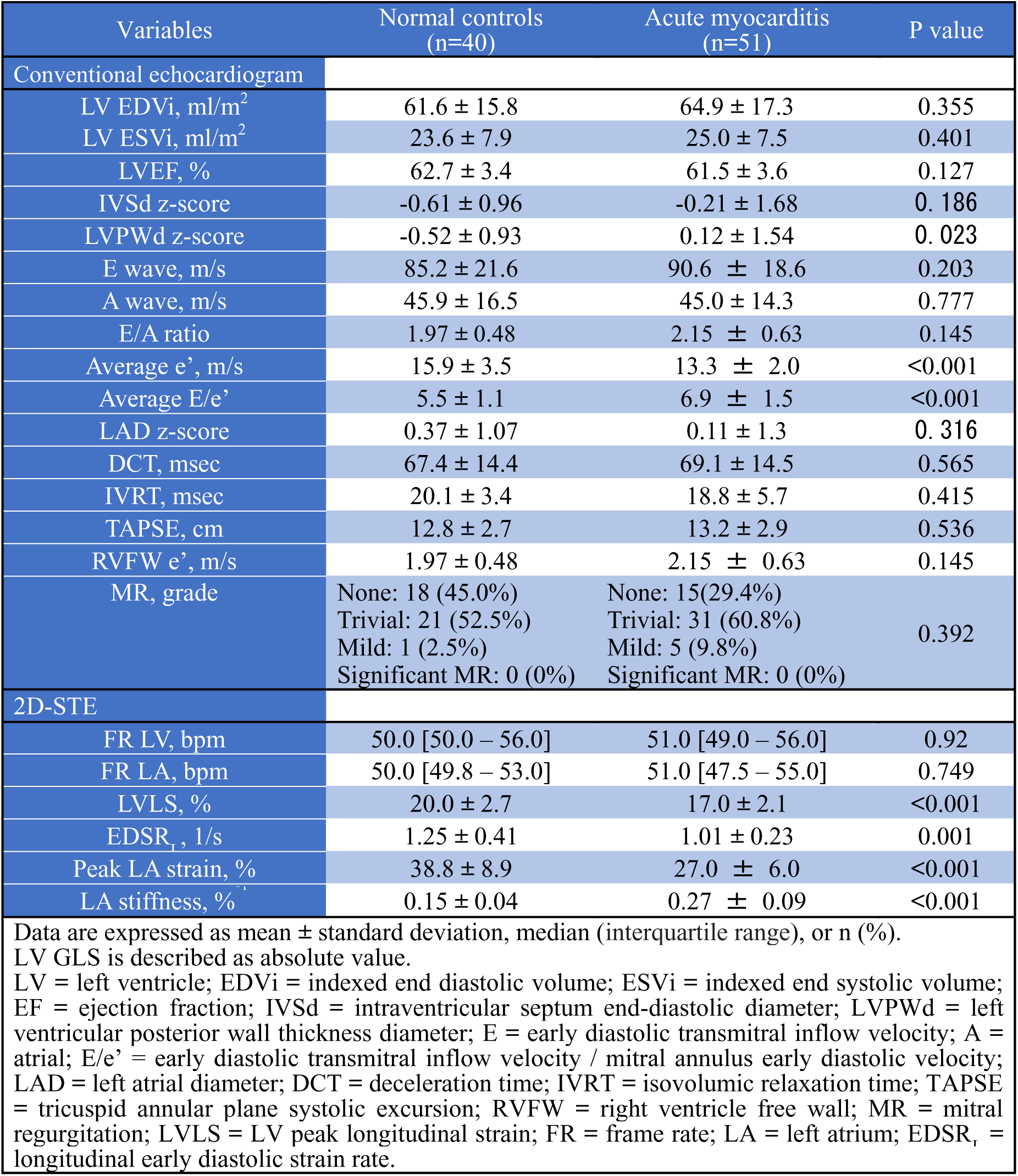
Echocardiographic features.

| Variables | Normal controls<br>(n=40) | Acute myocarditis<br>(n=51) | P value |
| --- | --- | --- | --- |
| Conventional echocardiogram |  |  |  |
| LV EDVi, ml/m<sup>2</sup> | 61.6 ± 15.8 | 64.9 ± 17.3 | 0.355 |
| LV ESVi, ml/m<sup>2</sup> | 23.6 ± 7.9 | 25.0 ± 7.5 | 0.401 |
| LVEF, % | 62.7 ± 3.4 | 61.5 ± 3.6 | 0.127 |
| IVSd z-score | -0.61 ± 0.96 | -0.21 ± 1.68 | 0.186 |
| LVPWd z-score | -0.52 ± 0.93 | 0.12 ± 1.54 | 0.023 |
| E wave, m/s | 85.2 ± 21.6 | 90.6 ± 18.6 | 0.203 |
| A wave, m/s | 45.9 ± 16.5 | 45.0 ± 14.3 | 0.777 |
| E/A ratio | 1.97 ± 0.48 | 2.15 ± 0.63 | 0.145 |
| Average e', m/s | 15.9 ± 3.5 | 13.3 ± 2.0 | <0.001 |
| Average E/e' | 5.5 ± 1.1 | 6.9 ± 1.5 | <0.001 |
| LAD z-score | 0.37 ± 1.07 | 0.11 ± 1.3 | 0.316 |
| DCT, msec | 67.4 ± 14.4 | 69.1 ± 14.5 | 0.565 |
| IVRT, msec | 20.1 ± 3.4 | 18.8 ± 5.7 | 0.415 |
| TAPSE, cm | 12.8 ± 2.7 | 13.2 ± 2.9 | 0.536 |
| RVFW e', m/s | 1.97 ± 0.48 | 2.15 ± 0.63 | 0.145 |
| MR, grade | None: 18 (45.0%)<br>Trivial: 21 (52.5%)<br>Mild: 1 (2.5%)<br>Significant MR: 0 (0%) | None: 15 (29.4%)<br>Trivial: 31 (60.8%)<br>Mild: 5 (9.8%)<br>Significant MR: 0 (0%) | 0.392 |
| 2D-STE |  |  |  |
| FR LV, bpm | 50.0 [50.0 – 56.0] | 51.0 [49.0 – 56.0] | 0.92 |
| FR LA, bpm | 50.0 [49.8 – 53.0] | 51.0 [47.5 – 55.0] | 0.749 |
| LVLS, % | 20.0 ± 2.7 | 17.0 ± 2.1 | <0.001 |
| EDSR<sub>1</sub>, 1/s | 1.25 ± 0.41 | 1.01 ± 0.23 | 0.001 |
| Peak LA strain, % | 38.8 ± 8.9 | 27.0 ± 6.0 | <0.001 |
| LA stiffness, % | 0.15 ± 0.04 | 0.27 ± 0.09 | <0.001 |
| Data are expressed as mean ± standard deviation, median (interquartile range), or n (%).<br>LV GLS is described as absolute value.<br>LV = left ventricle; EDVi = indexed end diastolic volume; ESVi = indexed end systolic volume;<br>EF = ejection fraction; IVSd = intraventricular septum end-diastolic diameter; LVPWd = left<br>ventricular posterior wall thickness diameter; E = early diastolic transmitral inflow velocity; A =<br>atrial; E/e' = early diastolic transmitral inflow velocity / mitral annulus early diastolic velocity;<br>LAD = left atrial diameter; DCT = deceleration time; IVRT = isovolumic relaxation time; TAPSE<br>= tricuspid annular plane systolic excursion; RVFW = right ventricle free wall; MR = mitral<br>regurgitation; LVLS = LV peak longitudinal strain; FR = frame rate; LA = left atrium; EDSR<sub>1</sub> =<br>longitudinal early diastolic strain rate. |  |  |  |

#### Echocardiographic comparisons between acute myocarditis and normal controls

Conventional echocardiogram showed no significant difference in indexed LV end-diastolic volume (LVEDVi) and end-systolic volume (LVESVi) (Table2). Patients with acute myocarditis had increased thickness of the LV posterior wall (LVPWd) but not of the septum. Diastolic impairment in the acute myocarditis group was represented by both lower average e’ and higher average E/e’, with no significant differences noted in other conventional and nonconventional indices. There was also no significant difference in the degree of mitral valve regurgitation between two groups.

Regarding 2D-STE parameters, patients with acute myocarditis had significantly lower LVLS compared with controls (mean LVLS (absolute value) 17.0 ± 2.1 vs. 20.0 ± 2.7 %, p<0.001). LA stiffness was significantly increased in acute myocarditis patients, accompanied by a reduced peak LA strain (mean LA stiffness 0.27 ± 0.09 vs. 0.15 ± 0.04 %^-1^; mean peak LA strain 27.0 ± 6.0 vs. 38.8 ± 8.9 %, p<0.001, respectively, Figure 2B).

**Figure 2.**
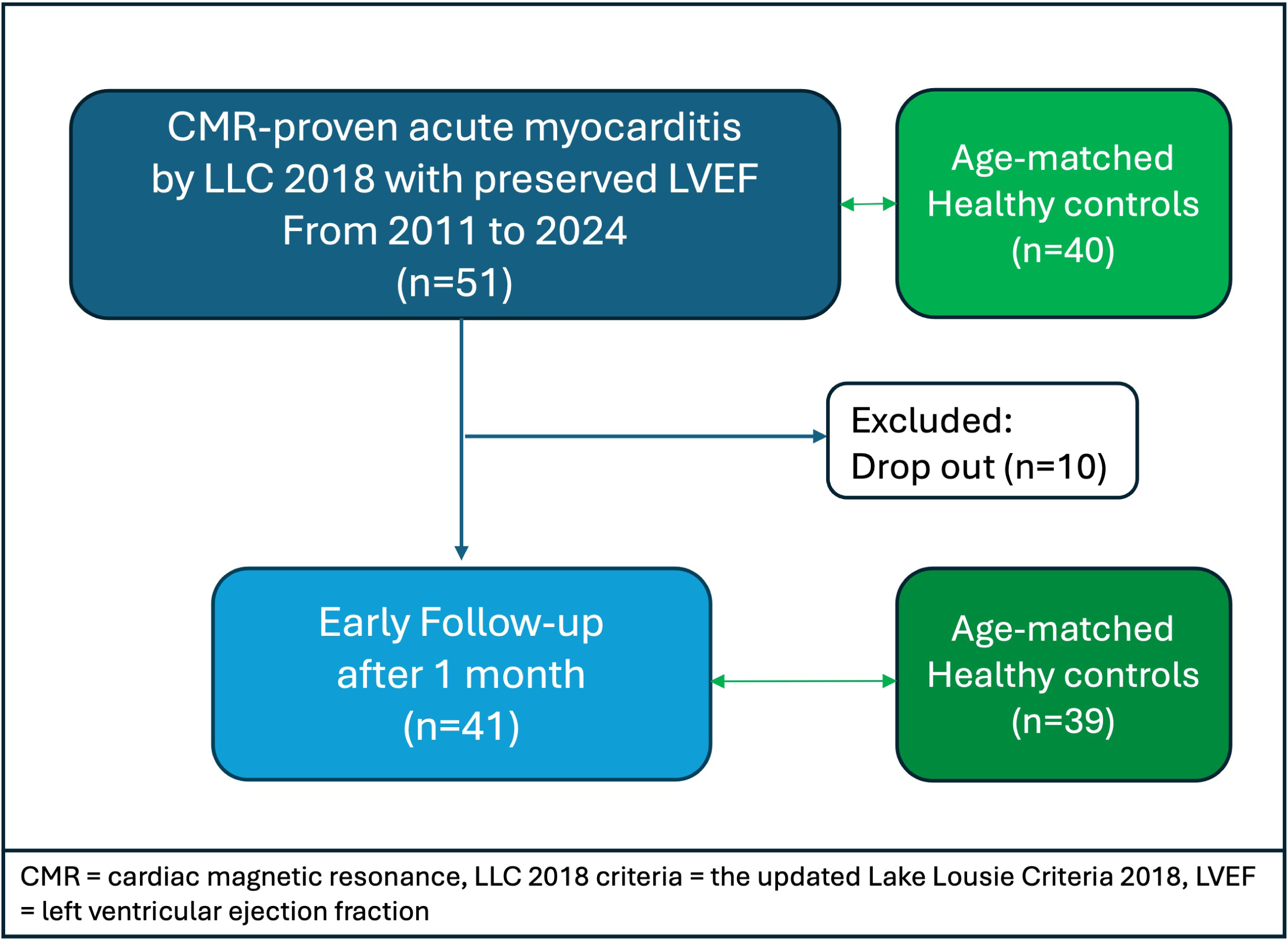
A, B. Left atrial two-dimensional speckle-tracking echocardiography and time strain curve. Example of left atrial (LA) strain imaging of the four-chamber view derived from Tomtec software (A), showing the different time strain curve between patients with acute myocarditis and controls (B).

#### Diagnostic performance of 2D-STE parameters in identifying acute myocarditis with preserved LVEF

LA stiffness showed the highest AUC of 0.944, with a sensitivity of 88.2 % and specificity of 92.5% at a cut-off value of 0.20 %^-1^, followed by peak LA strain (AUC: 0.897 [95% CI: 0.835 – 0.959]) and LVLS (AUC: 0.824 [95% CI: 0.733 – 0.914], Figure 3). The AUC of LA stiffness surpassed that of other indices apart from peak LA strain (Table 3). From our *Composite Score* = |*LVLS*(%)| − *LA stiffness*(%^−1^) × 100, a binary cut point was established at the Youden index, such that a composite score < 2.1 classified patients as likely acute myocarditis. This score had an AUC of 0.957 (95% CI: 0.914 – 1.000), with a 98% sensitivity and 87.5% specificity. LA stiffness and LVLS also showed an independent diagnostic power in multivariable regression analysis (LA stiffness, per 0.01 %^-1^ increase; OR 1.58 [95% CI: 1.32 – 1.89], LVLS, per 1% decrease; OR 1.72 [95% CI 1.36 – 2.17], p<0.001, respectively).

**Figure 3.**
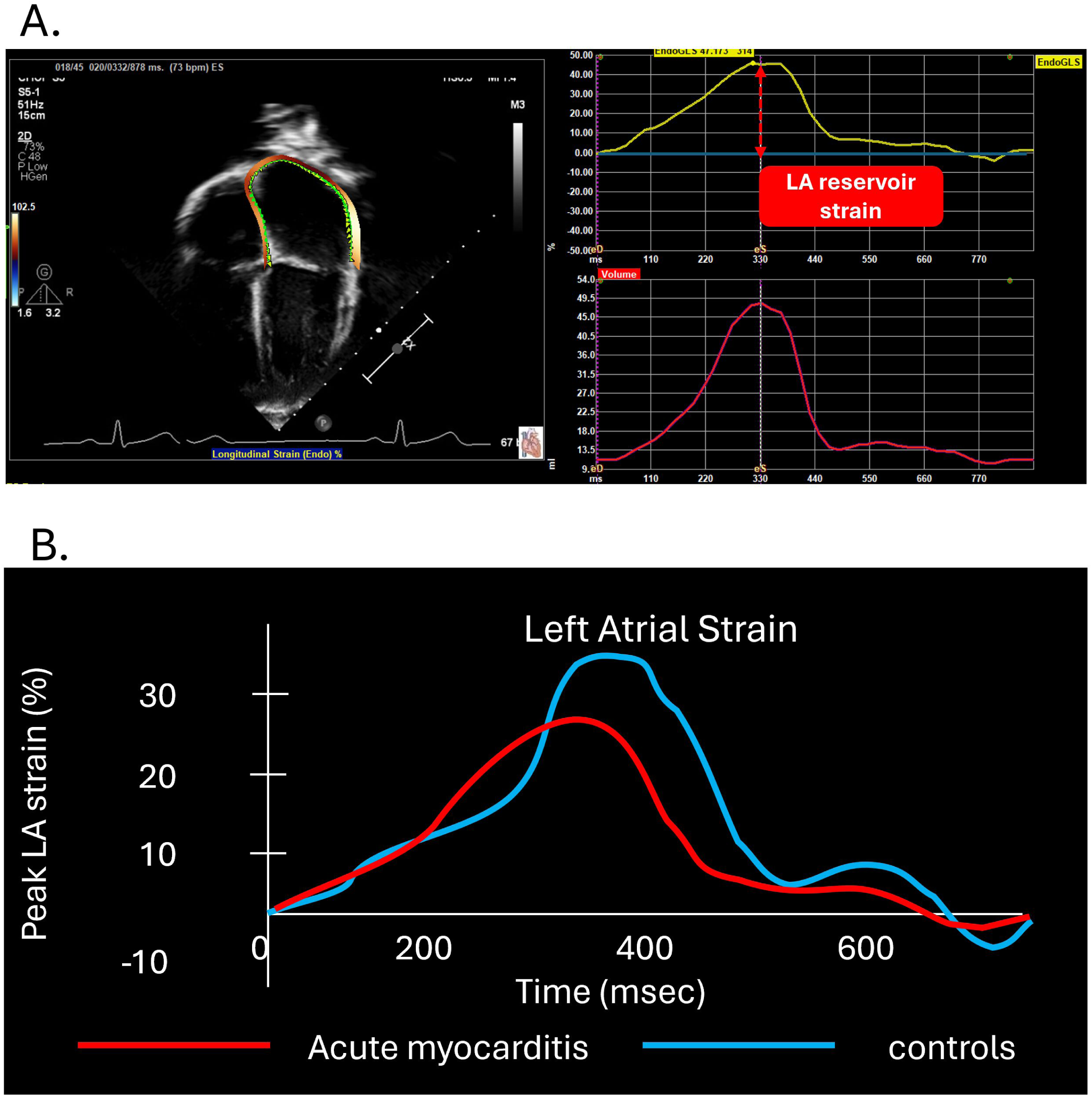
Diagnostic performance of LA stiffness in acute myocarditis patients with preserved LVEF. The diagnostic performance of LA stiffness was higher than that of other echocardiographic indices, with an area under the curve (AUC) of 0.944. When LV peak longitudinal strain (LVLS) was incorporated with LA stiffness into a single combined index, the AUC increased further to 0.957 (sensitivity 98.0%, specificity 87.5%; cut-off 2.1). CI = confidence interval

**Table 3.** Diagnostic performance of echocardiographic parameters in acute myocarditis patients with preserved LVEF.

| variables | AUC | 95% CI | threshold | Sensitivity | specificity | *p value* | DeLong test |
| --- | --- | --- | --- | --- | --- | --- | --- |
| LA stiffness, %<sup>-1</sup> | 0.944 | 0.901 – 0.987 | 0.20 | 88.2 % | 92.5 % | <0.001 | reference |
| Peak LA strain, % | 0.897 | 0.835 – 0.959 | 28.6 | 68.6 % | 97.5 % | <0.001 | 0.172 |
| LVLS, % | 0.824 | 0.733 – 0.914 | 18.8 | 78.4 % | 72.5 % | <0.001 | 0.008 |
| EDSR<sub>L</sub>, 1/s | 0.683 | 0.560 – 0.807 | 1.13 | 78.4 % | 65.0 % | 0.002 | <0.001 |
| Average E/e' | 0.787 | 0.694 – 0.880 | 5.86 | 76.5 % | 72.5 % | <0.001 | <0.001 |
| LAD z score | 0.563 | 0.444 – 0.682 | 0.32 | 60.8 % | 52.5 % | 0.314 | <0.001 |
| Composite Score* | 0.957 | 0.914 – 1.000 | 2.1 | 98.0 % | 87.5 % | <0.001 | 0.422 |

LV GLS and GCS are described as absolute values. \*composite model: |LVLS| - LA stiffness\*100
AUC = areas under the curve; CI = confidence interval; LA = left atrium; LVLS = left ventricular peak longitudinal strain; EDSR<sub>L</sub> = longitudinal early diastolic strain rate; E/e' = early diastolic transmitral inflow velocity / mitral annulus early diastolic velocity; LAD = left atrial diameter.

#### Correlation between LA stiffness and laboratory markers of myocarditis

LA stiffness showed the best correlation with peak BNP values during the acute phase (r = 0.657, p<0.001, Figure 4) among other echocardiographic indices (Supplemental table 1). Despite elevated TnI levels in all patients, there were no significant correlations between echocardiographic parameters, including LA stiffness, with peak TnI levels.

**Figure 4.**
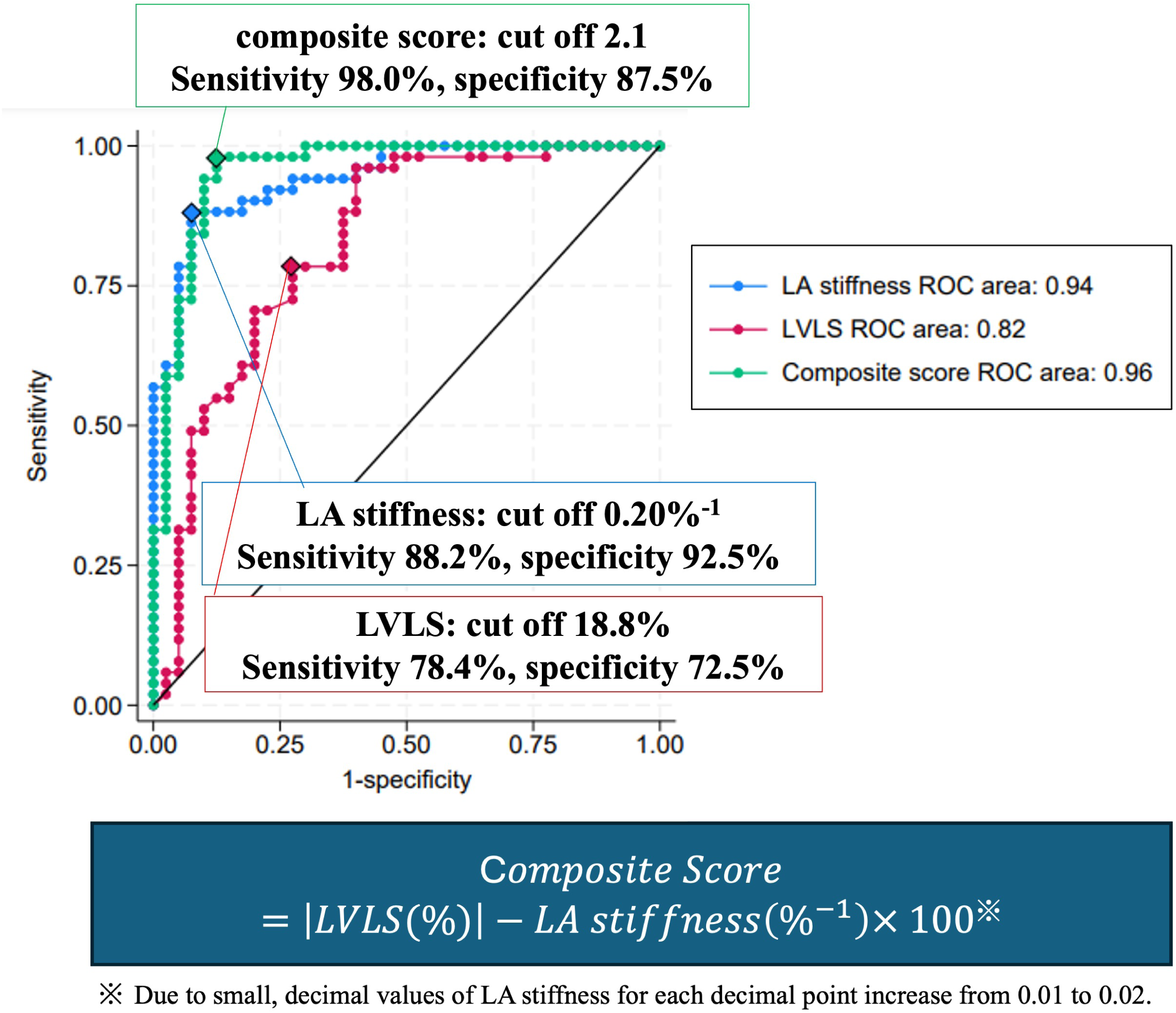
Correlation among LA stiffness, LV peak longitudinal strain, and biomarkers. The left atrial (LA) stiffness showed the strongest correlation with brain-type natriuretic peptide (BNP), while troponin I showed no correlation. Abbreviations are as defined in Figure 3 and 4

### Follow-up cohort

#### Changes from acute phase to follow-up stage in pediatric myocarditis

In our cohort, only two patients with preserved-LVEF experienced life-threatening arrythmia during admission and subsequently implantable cardiovascular defibrillators were placed. The other patients had no adverse cardiac events during the follow-up period (median 9.6 [3.1 – 23.4] months). Table 4 presents the demographic data and echocardiographic characteristics of patients with myocarditis during follow-up. Of the 51 patients, 41 (80.4%) underwent follow-up echocardiographic evaluation.

**Table 4.**
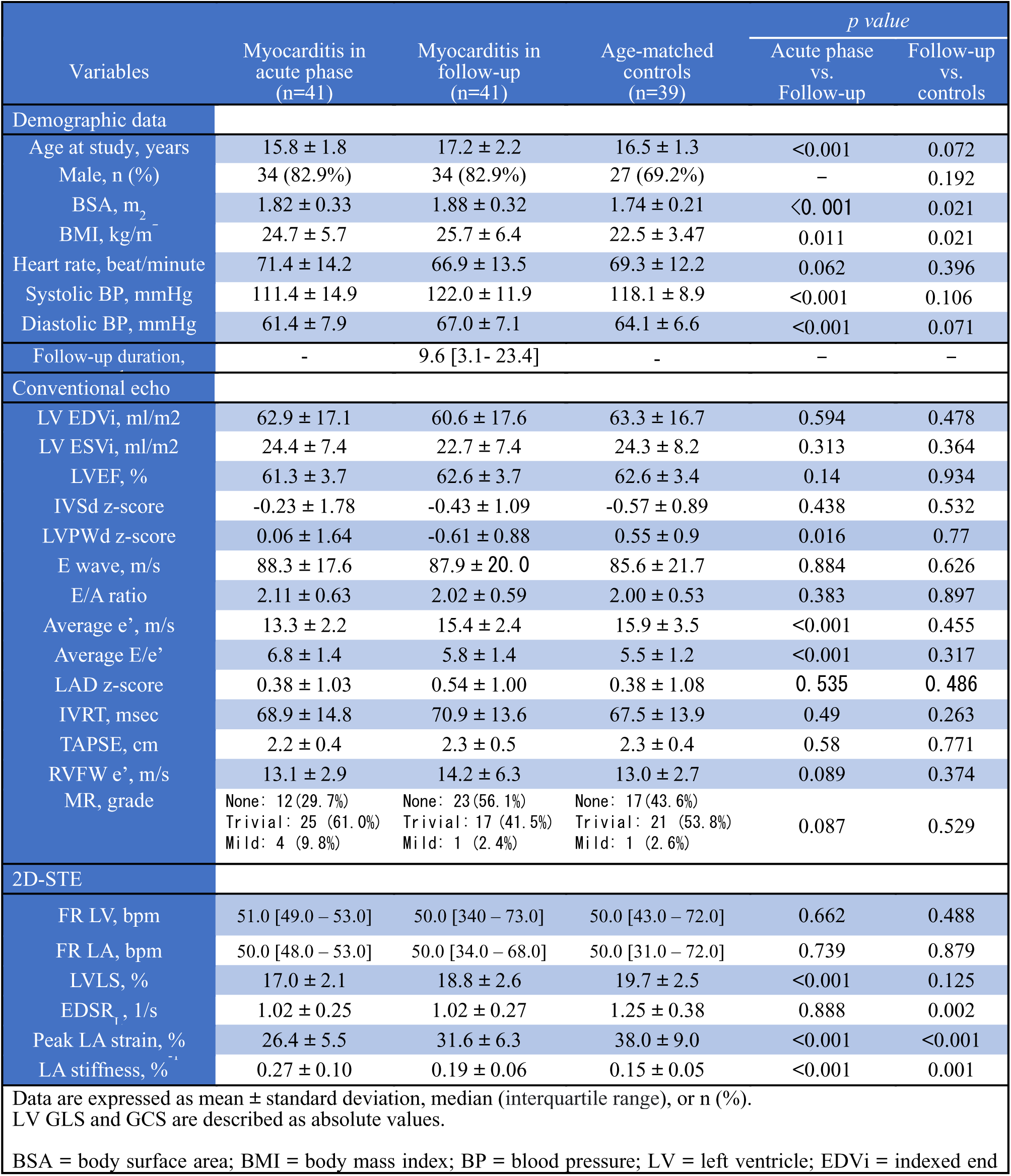
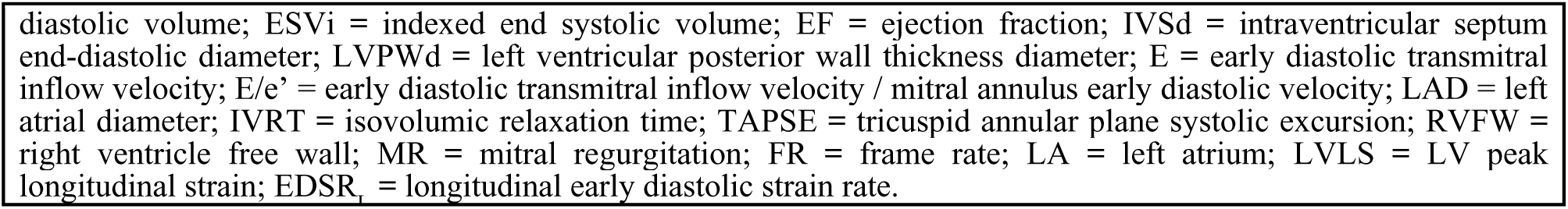
Echocardiographic features between the acute phase and the follow-up in patients with acute myocarditis compared with age-matched controls.

| Variables | Myocarditis in acute phase (n=41) | Myocarditis in follow-up (n=41) | Age-matched controls (n=39) | *p value* |  |
| --- | --- | --- | --- | --- | --- |
|  |  |  |  | Acute phase vs. Follow-up | Follow-up vs. controls |
| Demographic data |  |  |  |  |  |
| Age at study, years | 15.8 ± 1.8 | 17.2 ± 2.2 | 16.5 ± 1.3 | <0.001 | 0.072 |
| Male, n (%) | 34 (82.9%) | 34 (82.9%) | 27 (69.2%) | – | 0.192 |
| BSA, m<sub>2</sub> | 1.82 ± 0.33 | 1.88 ± 0.32 | 1.74 ± 0.21 | <0.001 | 0.021 |
| BMI, kg/m<sup>2</sup> | 24.7 ± 5.7 | 25.7 ± 6.4 | 22.5 ± 3.47 | 0.011 | 0.021 |
| Heart rate, beat/minute | 71.4 ± 14.2 | 66.9 ± 13.5 | 69.3 ± 12.2 | 0.062 | 0.396 |
| Systolic BP, mmHg | 111.4 ± 14.9 | 122.0 ± 11.9 | 118.1 ± 8.9 | <0.001 | 0.106 |
| Diastolic BP, mmHg | 61.4 ± 7.9 | 67.0 ± 7.1 | 64.1 ± 6.6 | <0.001 | 0.071 |
| Follow-up duration, | - | 9.6 [3.1- 23.4] | - | - | - |
| Conventional echo |  |  |  |  |  |
| LV EDVi, ml/m2 | 62.9 ± 17.1 | 60.6 ± 17.6 | 63.3 ± 16.7 | 0.594 | 0.478 |
| LV ESVi, ml/m2 | 24.4 ± 7.4 | 22.7 ± 7.4 | 24.3 ± 8.2 | 0.313 | 0.364 |
| LVEF, % | 61.3 ± 3.7 | 62.6 ± 3.7 | 62.6 ± 3.4 | 0.14 | 0.934 |
| IVSd z-score | -0.23 ± 1.78 | -0.43 ± 1.09 | -0.57 ± 0.89 | 0.438 | 0.532 |
| LVPWd z-score | 0.06 ± 1.64 | -0.61 ± 0.88 | 0.55 ± 0.9 | 0.016 | 0.77 |
| E wave, m/s | 88.3 ± 17.6 | 87.9 ± 20.0 | 85.6 ± 21.7 | 0.884 | 0.626 |
| E/A ratio | 2.11 ± 0.63 | 2.02 ± 0.59 | 2.00 ± 0.53 | 0.383 | 0.897 |
| Average e', m/s | 13.3 ± 2.2 | 15.4 ± 2.4 | 15.9 ± 3.5 | <0.001 | 0.455 |
| Average E/e' | 6.8 ± 1.4 | 5.8 ± 1.4 | 5.5 ± 1.2 | <0.001 | 0.317 |
| LAD z-score | 0.38 ± 1.03 | 0.54 ± 1.00 | 0.38 ± 1.08 | 0.535 | 0.486 |
| IVRT, msec | 68.9 ± 14.8 | 70.9 ± 13.6 | 67.5 ± 13.9 | 0.49 | 0.263 |
| TAPSE, cm | 2.2 ± 0.4 | 2.3 ± 0.5 | 2.3 ± 0.4 | 0.58 | 0.771 |
| RVFW e', m/s | 13.1 ± 2.9 | 14.2 ± 6.3 | 13.0 ± 2.7 | 0.089 | 0.374 |
| MR, grade | None: 12 (29.7%)<br>Trivial: 25 (61.0%)<br>Mild: 4 (9.8%) | None: 23 (56.1%)<br>Trivial: 17 (41.5%)<br>Mild: 1 (2.4%) | None: 17 (43.6%)<br>Trivial: 21 (53.8%)<br>Mild: 1 (2.6%) | 0.087 | 0.529 |
| 2D-STE |  |  |  |  |  |
| FR LV, bpm | 51.0 [49.0 – 53.0] | 50.0 [340 – 73.0] | 50.0 [43.0 – 72.0] | 0.662 | 0.488 |
| FR LA, bpm | 50.0 [48.0 – 53.0] | 50.0 [34.0 – 68.0] | 50.0 [31.0 – 72.0] | 0.739 | 0.879 |
| LVLS, % | 17.0 ± 2.1 | 18.8 ± 2.6 | 19.7 ± 2.5 | <0.001 | 0.125 |
| EDSR<sub>1</sub>, 1/s | 1.02 ± 0.25 | 1.02 ± 0.27 | 1.25 ± 0.38 | 0.888 | 0.002 |
| Peak LA strain, % | 26.4 ± 5.5 | 31.6 ± 6.3 | 38.0 ± 9.0 | <0.001 | <0.001 |
| LA stiffness, %<sup>-1</sup> | 0.27 ± 0.10 | 0.19 ± 0.06 | 0.15 ± 0.05 | <0.001 | 0.001 |
| Data are expressed as mean ± standard deviation, median (interquartile range), or n (%).<br>LV GLS and GCS are described as absolute values. |  |  |  |  |  |
| BSA = body surface area; BMI = body mass index; BP = blood pressure; LV = left ventricle; EDVi = indexed end |  |  |  |  |  |

diastolic volume; ESV<sub>i</sub> = indexed end systolic volume; EF = ejection fraction; IVS<sub>d</sub> = intraventricular septum end-diastolic diameter; LVPW<sub>d</sub> = left ventricular posterior wall thickness diameter; E = early diastolic transmitral inflow velocity; E/e' = early diastolic transmitral inflow velocity / mitral annulus early diastolic velocity; LAD = left atrial diameter; IVRT = isovolumic relaxation time; TAPSE = tricuspid annular plane systolic excursion; RVFW = right ventricle free wall; MR = mitral regurgitation; FR = frame rate; LA = left atrium; LVLS = LV peak longitudinal strain; EDSR<sub>e</sub> = longitudinal early diastolic strain rate.

Figure 5 and Table 4 show that LA stiffness showed improvement during follow-up (from 0.27 ± 0.10 to 0.19 ± 0.06 %^-1^, p < 0.001), although it remained abnormal compared to controls (0.19 ± 0.06 vs. 0.15 ± 0.05 %^-1^, p = 0.001). Similarly, peak LA strain improved at follow-up, but remained lower compared to controls. In contrast, conventional diastolic parameters and LVLS normalized during follow-up (Table 4).

**Figure 5.**
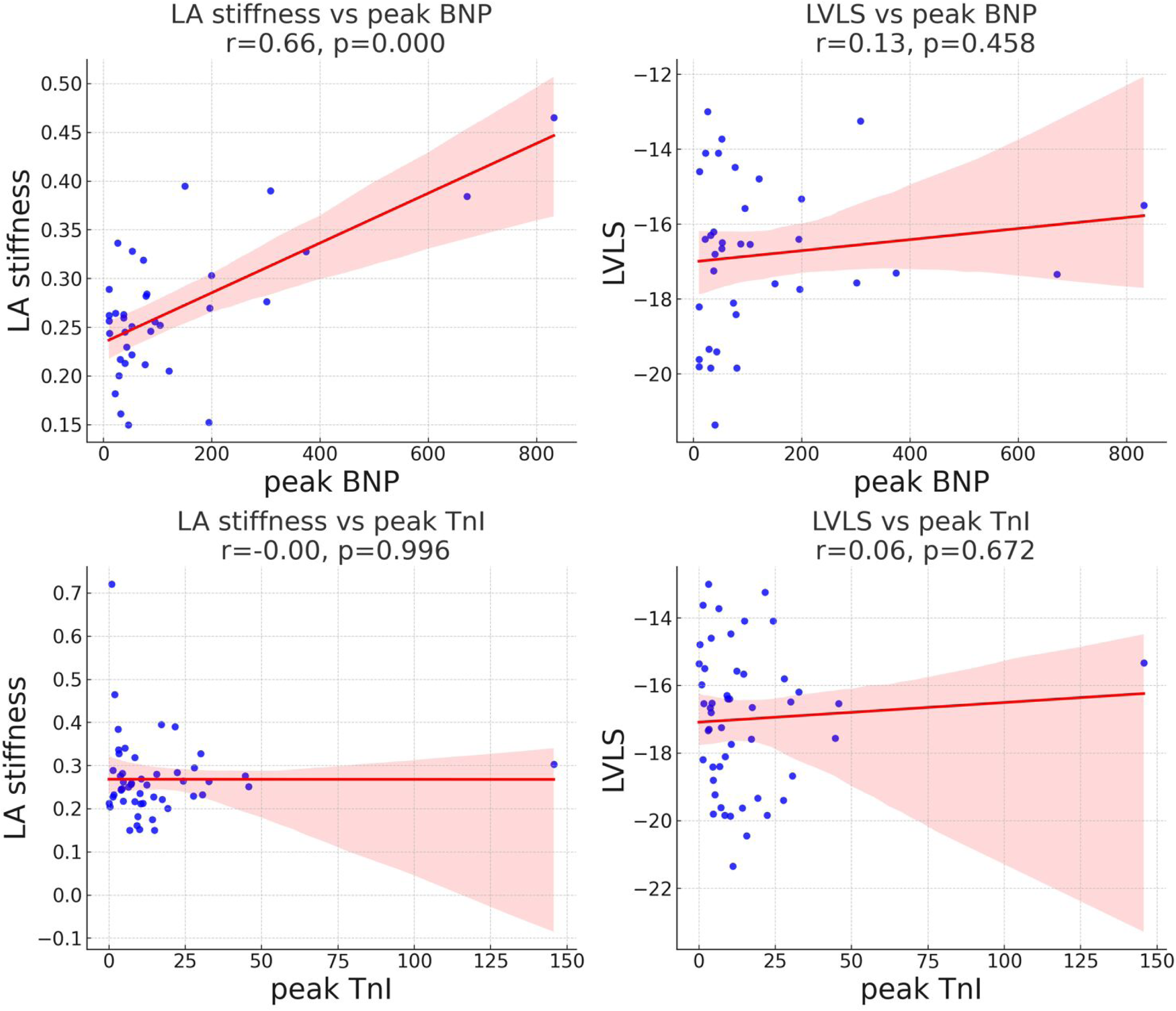
Echocardiographic fluctuations between the acute phase and follow-up in patients with acute myocarditis. Pediatric acute myocarditis showed significant improvement in LA stiffness, peak LA strain, and LVLS during follow-up. However, impaired LA stiffness and LA strain persisted when compared to controls, while LVLS normalized.

#### Reproducibility analysis

Our measurement of LA stiffness and LA strain showed excellent intra- and inter-observer reliability (LA stiffness: ICCs, 0.95; bias 0.0002%^-1^ [95% LOA, -0.049 to 0.049]; and 0.93; bias 0.005%^-1^ [95% LOA, -0.065 to 0.049], and peak LA strain: ICCs, 0.92; bias 1.03% [95% LOA, -6.57 to 8.63]; and 0.95; bias -1.03% [95% LOA, -7.59 to 5.53], respectively, Supplemental Table 2). These results indicated reliability within the same observer and good agreement between the two observers (Supplemental Figure 1).

## Discussion

The emphasis of our study was on a selective cohort of CMR-proven myocarditis with preserved-LVEF. We found that 1) noninvasive LA stiffness, derived from 2D-STE, demonstrated the highest diagnostic accuracy in differentiating myocarditis patients with preserved EF from healthy controls, and 2) this novel measure impacted the longitudinal assessment of diastolic dysfunction in this population. Notably, LA stiffness was impaired even in the early follow-up period in pediatric patients with acute myocarditis and preserved-LVEF. Additionally, among all echocardiographic parameters, LA stiffness showed the strongest correlation with BNP levels.

### Diastolic dysfunction in pediatric patients with acute myocarditis

Impaired diastolic function was observed in adult patients with acute myocarditis from the acute phase through mid-term follow-up (54–78 months).^7^ Those adults with myocarditis showed reduced LA reservoir and conduit strain derived by CMR, despite preserved-LVEF, consistent with impairment of conventional diastolic indices.^15,17^ Similarly, in children, our results exhibited reduced LA reservoir strain and increased LA stiffness in acute myocarditis patients with preserved-LVEF, accompanied by a higher E/e’.

Unlike adults, noninvasive assessment of diastolic dysfunction has not been well-established in children.^22^ Previous reports have shown that the adult diagnostic algorithms of the ASE incorrectly classified up to 30% of children with overt, often severe cardiomyopathy as having normal diastolic function, even when pediatric reference values were applied in place of adult cutoffs. In their study, LA volume and e’ were often discordant even in well-established cardiomyopathy, frequently leading to misclassification of patients with significant LV diastolic impairment as normal and vice versa.^23^ Sasaki et al. similarly assessed the applicability of published guidelines for evaluating LV diastolic dysfunction in children with restrictive cardiomyopathy (RCM), finding substantial overlap in E/e’ and e’ values between controls and affected patients. As a result, no reliable cutoff could be established for identifying RCM.^24^ Both studies have concluded that additional indices are needed for noninvasive assessment of LV diastolic dysfunction in children.

To address these limitations, we focused on LA stiffness, as a novel noninvasive echocardiographic parameter. Initially described in adults with severe LV hypertrophy undergoing cardiac catheterization, LA stiffness was found to correlate significantly with PCWP, highlighting its potential for assessing LV diastolic dysfunction.^8^ In children, our previous studies demonstrated a strong correlation of both LA stiffness and LA strain with PCWP, as well as superior reliability in differentiating abnormal PCWP (>12 mm Hg) compared to E/e’.^13^ More recently, we have demonstrated that in pediatric patients with HCM - characterized by preserved LVEF and diastolic dysfunction, LA stiffness was markedly elevated and exhibited the strongest correlation with BNP among all diastolic indices.^14^ Consistently, the present study also found that LA stiffness had the strongest correlation with BNP in pediatric acute myocarditis.

Furthermore, this study showed that impaired LA stiffness persisted during early follow-up, despite improvement and normalization of conventional diastolic parameters. Escher et al. reported that a slow but progressive profibrotic process in adult patients with myocarditis is associated with the persistent or development of diastolic dysfunction, resulting in increased risk of mortality and morbidity.^7^ Therefore, LA stiffness may serve as a sensitive, noninvasive biomarker reflecting diastolic dysfunction, providing valuable information for appropriate management during long-term follow-up, even after the acute phase in children.

### Diagnostic reliability of LA stiffness in acute myocarditis with preserved EF

Because of the tissue characterization techniques in CMR, it has been utilized as a gold standard for the diagnosis of acute myocarditis using the LLC, updated in 2018.^3^ CMR can evaluate the entire myocardium as opposed to endomyocardial biopsy, which typically is obtained from the right ventricular septum and may miss regions of inflammation in other regions of the heart. One of the first publications of CMR findings and tissue characterization in myocarditis was in children in 1991 which compared these findings favorably to endomyocardial biopsy.^3^ Several studies have reported that 2D-STE could potentially provide additional diagnostic and prognostic data, especially in patients with preserved-EF, where conventional indices might not fully capture the extent of myocardial involvement.^25^ Specifically, LA strain parameters emerged as crucial indicators of cardiac function in conditions such as hypertrophic cardiomyopathy and acute myocarditis.^13,14,26^ This is particularly relevant in myocarditis, where atrial modification and diastolic impairment can occur even when LVEF is preserved.^27^ Zhang et al. highlighted the importance of LA deformation analysis in identifying adult acute myocarditis. In this report, LA strain indices proved to be more diagnostically valuable, than strain measurements from other chambers, when LVEF was preserved.^9^ Dick et al. also demonstrated that LA strain parameters showed the highest diagnostic accuracy, with a specificity of 92.9%.^16^ LA stiffness provides an additional diagnostic advantage, as our previous report demonstrated that LA stiffness provides a better understanding of atrioventricular coupling than LA strain alone.^13^ In the present study, when LVLS was combined with LA stiffness into a single model, diagnostic accuracy was the highest among all echocardiographic parameters. Moreover, it is noteworthy that LA stiffness had independent power for differentiating acute myocarditis in multivariable analysis. Therefore, LA stiffness (cutoff value > 0.2%^-1^) holds strong diagnostic potential for the early detection of acute myocarditis in pediatric patients, consistent with the similar cutoff value, approximately 0.25-0.27%^-1^, previously reported for identifying diastolic impairment in children.^13,28^ This index may allow physicians to identify and closely monitor suspected cases of myocarditis in the clinical setting, even before a definitive diagnosis is made using CMR. By using combined deformation analyses of both LV and LA, the diagnostic value is further boosted. With an AUC of 0.957 (sensitivity 98.0%, specificity 87.5%), this combined index has a robust capability for diagnosing acute myocarditis in children. Additionally, these parameters can help detect persistent diastolic dysfunction during follow-up visits. The value of 2D-STE may be particularly relevant in many global settings where access to CMR is limited, offering a less invasive alternative that does not require sedation in children. Interestingly, LA stiffness has garnered attention not only as a diagnostic marker but also for its prognostic significance in patients with preserved LVEF, both in adults and children.^11,26^ The follow-up period for the present study is short and future longitudinal studies with longer follow-up periods may further elucidate the prognostic utility of LA stiffness in predicting adverse outcomes over extended periods of time in pediatric patients with acute myocarditis.

### Study Limitations

We utilized the four-chamber LV strain exclusively, as two- and three-chamber views were unavailable for several patients. This study covers a period exceeding 14 years, during which the two- and three-chamber views were not routinely acquired in our laboratory. However, prior studies have demonstrated that the apical four-chamber longitudinal strain is a universally applicable index with excellent intra- and inter-observer reliability for evaluating systolic function, showing a strong correlation with EF.^29^ Therefore, we consider this method acceptable for assessing LV systolic function in our study. Additionally, we focused solely on the peak LA strain value to evaluate LA deformation. Unlike adult studies, detecting the precise incisura just before atrial contraction in LA time-strain-curve is often challenging in pediatric patients (where magnitude of LA contraction is less than adults). To minimize the risk of observer bias, we excluded both conduit and contractile strain measurements. Finally, our follow-up period was relatively short, and further longitudinal studies are needed to describe the trajectory of LA stiffness in patients with preserved EF, which was beyond the scope of the present manuscript.

## Conclusion

In this study we evaluated a selective cohort of patients with CMR-proven myocarditis and preserved LVEF. LA stiffness is a novel index that has been utilized to predict elevated PCWP in children. An optimal cutoff of 0.2%^-1^ of LA stiffness may offer incremental value as a robust echocardiographic marker for consolidating the diagnosis of acute myocarditis in conjunction with CMR. By combining atrio-ventricular indices into a combined index, the diagnostic ability for detecting subtle myocarditis was even further boosted. LA stiffness also proved to be reliable for assessment of LV diastolic dysfunction during the later follow-up stages of myocarditis, when LVEF was preserved.

## Acknowledgement

None

## Data availability

The data underlying this article will be shared upon reasonable request to the corresponding author.

## Supplemental Material

Supplemental Tables S1–S2

Figures S1

## Abbreviation

CMR: cardiac magnetic resonance imaging
2D-STE: two-dimensional speckle-tracking echocardiography
HFpEF: heart failure with preserved ejection fraction
PCWP: pulmonary capillary wedge pressure
LLC: updated Lake Louise criteria
TTE: transthoracic echocardiogram
BNP: brain natriuretic peptide
TnI: troponin I
IVRT: isovolumic relaxation time
LVLS: left ventricular peak longitudinal strain
LVCS: left ventricular peak circumferential strain
EDSR_L_: longitudinal early diastolic strain rate
LVPWd: left ventricular posterior wall diameter
MR: mitral valve regurgitation;

---

**Supplemental figure 1.**
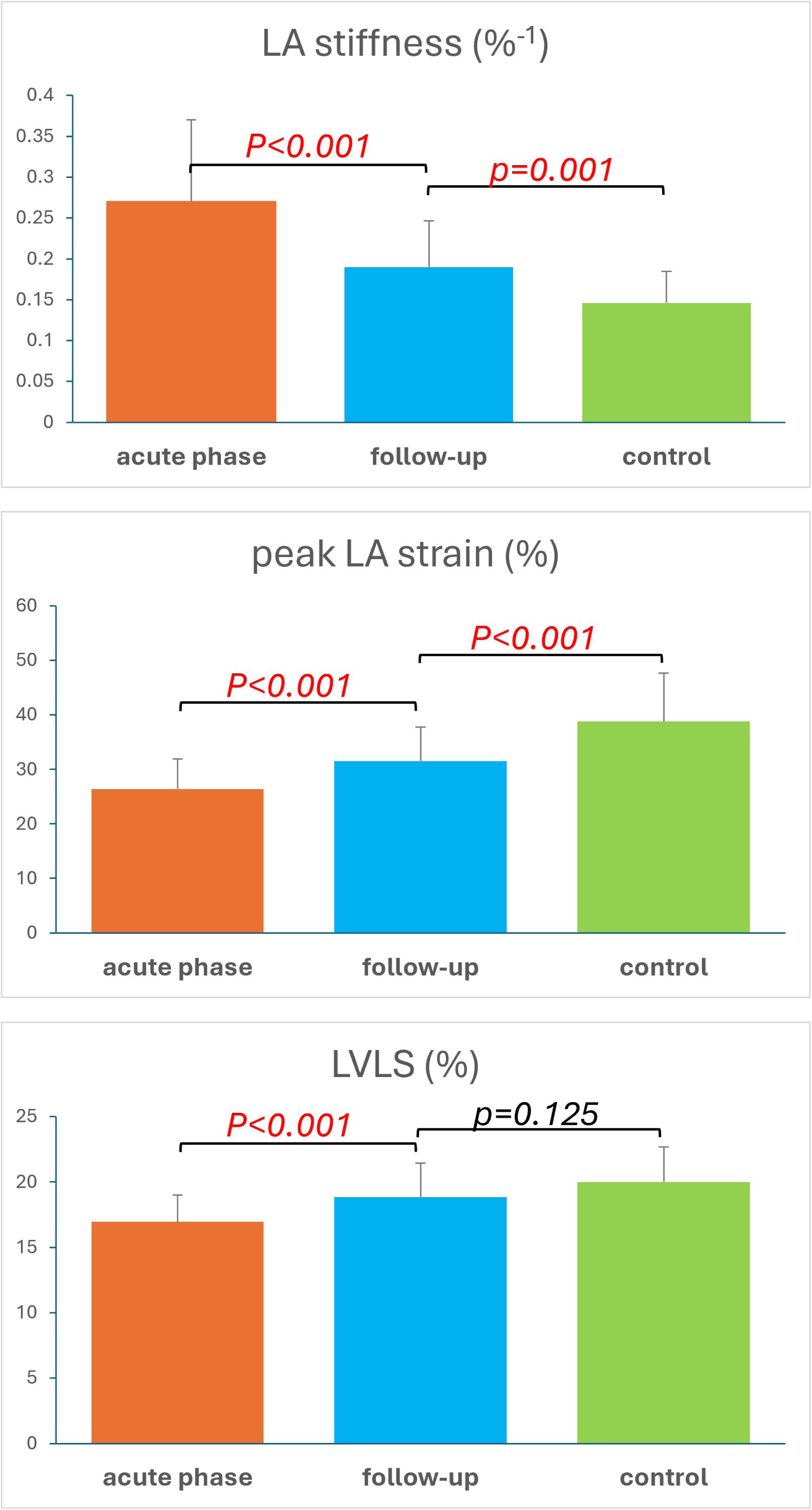
Reliability of left atrial stiffness expressed by intra- and inter-observer using Bland-Altman analysis.

## Central illustration

In our study, fifty-one pediatric patients with acute myocarditis and preserved EF were enrolled. We found that LA stiffness, when incorporated into composite score with LVLS, demonstrated superior diagnostic performance in this population and may provide incremental value for follow-up management. The high LA stiffness persisted from acute phase to early follow-up, and it showed a moderate correlation with BNP levels. Abbreviations are as defined in Figure 3 and 4

